# COVID-19 Testing and Vaccination among US Older Adults with Vision Impairment: The National Health and Aging Trends Study 2021

**DOI:** 10.1101/2023.08.23.23294511

**Authors:** Louay Almidani, Bonnielin K. Swenor, Joshua R. Ehrlich, Varshini Varadaraj

## Abstract

**Purpose:** To examine the associations between vision impairment (VI) and COVID-19 testing and vaccination services in older US adults.

**Methods:** This cross-sectional study assessed data from adults ≥65 years who participated in the National Health and Aging Trends Study (year 2021), a nationally representative sample of Medicare beneficiaries. Exposure: Distance VI (<20/40), near VI (<20/40), contrast sensitivity impairment (CSI) (<1.55 logCS), and any VI (distance, near, or CSI). Outcomes: Self-reported COVID-19 testing and vaccination.

**Results:** Of 2,822 older adults, the majority were female (weighted; 55%) and White (82%), and 32% had any VI. In fully-adjusted regression analyses, older adults with any VI had similar COVID-19 vaccination rates to adults without any VI (OR:0.77, 95% CI:0.54–1.09), but had lower odds of COVID-19 testing (OR:0.82, 95% CI:0.68–0.97). Older adults with distance (OR:0.47, 95% CI:0.22–0.99) and near (OR:0.68, 95% CI:0.47–0.99) VI were less likely to be vaccinated for COVID-19, while those with CSI were less likely to test for COVID-19 (OR:0.76, 95% CI:0.61– 0.95), as compared to peers without respective impairments. The remaining associations were not significant (p>.05).

**Conclusions and Relevance:** These findings highlight inequities in the COVID-19 pandemic response for people with vision disability and emphasize the need for equitable prioritization of accessibility of healthcare services for all Americans.

## 1. Introduction

COVID-19, stemming from the severe acute respiratory syndrome coronavirus 2 (SARS-CoV-2), is a significant contributor to morbidity and mortality, with the elderly population emerging as the most susceptible cohort.^1^ Further, older age is also often associated with an increased prevalence of vision impairment (VI).^2^ As a result, adults with VI may be at a higher risk of experiencing severe COVID-19 outcomes, underscoring the importance of preventive healthcare services, including vaccinations.

The development of the COVID-19 vaccine has caused a major shift during the pandemic, resulting in millions of lives saved globally. Watson et al (2022) estimated that vaccinations prevented over 14 million deaths from COVID-19 across 185 countries and territories.^3^ Nonetheless, the impact of vaccinations has been hindered by the inadequate vaccine accessibility in certain communities.^3^

The COVID-19 pandemic has had a disproportionate impact on people with disabilities, including those with VI.^4^ It has exacerbated existing barriers in accessing essential preventive healthcare services, such as vaccination and testing.^5^ Studies have shown that adults with VI are less likely to use preventive services,^6^ including lower vaccine initiation rates,^7^ and experience worse health outcomes than adults without VI.^8^ Additionally, individuals with VI may face challenges in navigating healthcare systems and accessing public health information, which could increase their risk of COVID-19 or other illnesses.^9^

Despite these associations, previous studies lacked generalizability or only assessed one aspect of vision. Vision is a complex process, and while distance visual acuity (the ability to see objects at a distance) is most frequently used in research studies as a measure of visual function,^10^ it only represents one component of visual function.^11^ Recent findings suggest that other aspects of vision, such as near visual acuity (the ability to see objects located close to the eyes, which is important for tasks like reading) and contrast sensitivity (the ability to perceive differences in shades and distinguish objects from their background), can also impact daily functioning.^12–14^

Accordingly, we utilized data from the National Health and Aging Trends Study (NHATS), a US population-based, nationally representative sample of Medicare beneficiaries, to assess the potential associations of three measures of vision — distance visual acuity (VA), near VA, and contrast sensitivity (CS) — with COVID-19 vaccination and testing in a national sample of older adults, as they are at risk of VI and worse COVID-19 outcomes.^4^

## 2. Materials and Methods

### 2.1 Study design and participants

This cross-sectional study presents data from the NHATS Round 11 (2021), a nationally representative sample of Medicare beneficiaries aged ≥65 years in the United States.^15^ In-home interviews and performance-based tests, including vision testing, were conducted annually from approximately June to November.^16^ Further details regarding survey sampling design have been previously described.^15,16^ Written informed consent was obtained from all participants or their proxies. This secondary analysis of publicly available data was acknowledged as exempt research by the institutional review board of the Johns Hopkins School of Medicine. The study adheres to the Strengthening the Reporting of Observational Studies in Epidemiology (STROBE) reporting guidelines for cross-sectional studies.

### 2.2 Vision Measures

In 2021, the NHATS introduced three objective vision tests (distance and near VA, and CS) that measured presenting binocular vision while wearing habitual correction (glasses or contacts). Tests were conducted via Ridgevue Vision tablet-based tests (ridgevue.com), which show good agreement with corresponding gold standard tests (ETDRS distance acuity, MNRead near acuity, and Pelli-Robson contrast sensitivity), and enable standardized monitoring to occur in a variety of settings, including at home or in rehabilitation facilities.^17,18^

For distance VA testing, participants sat five feet away from the tablet and were instructed to read five letters per screen with each subsequent screen displaying reduced letter size. For near VA testing, participants held the tablet at their usual reading distance and were instructed to read five letters per screen with each subsequent screen displaying smaller letters. For CS testing, participants were instructed to read two letters per screen, with the tone becoming lighter with each subsequent screen. Further details on how the tests were conducted have been described previously.^17,19^ Distance and near VA were calculated as the logarithm of the minimum angle of resolution (logMAR), with near VA accounting for reading distance. CS was measured in logCS units. We assessed vision on a continuous scale; distance and near VA (per 0.1 logMAR), and CS (per 0.1 logCS), and on a categorical scale; distance VI (<20/40), near VI (<20/40) – based on the US American Academy of Ophthalmology definitions,^20^ – and CS impairment (CSI) (<1.55), as previously defined.^21^ Any objective VI was defined as having VI in either distance, near, or CS, and was taken as the primary exposure.

### 2.3 Outcomes

We examined two sets of outcomes: COVID-19 vaccination and COVID-19 testing. Participants were considered to have been COVID-19 vaccinated or tested if they answered “yes” to any of the following questions respectively: (1) Have you been vaccinated for COVID-19? (2) Have you ever been tested for COVID-19? Outcomes were based on self-reports and were recorded in the NHATS dataset as binary variables (yes/no).

### 2.4 Covariates

Demographic characteristic covariates included age (categorized into five age intervals: 70-74, 75-79, 80-84, 85-89, and ≥90; continuous age is not provided in the publicly available NHATS data), gender (male, female), race (Non-Hispanic White, Non-Hispanic Black, Hispanic, and other), education (≤ high school, some college, and ≥ college graduate), living arrangements (alone or not alone), and number of comorbidities (0-1, 2, 3, and ≥4). Comorbidities include self-reported diagnoses of diabetes, hypertension, myocardial infarction, arthritis (osteoarthritis or rheumatoid arthritis), osteoporosis, stroke, lung disease (such as emphysema, asthma, or chronic bronchitis), cancer, and hip fracture. Covariates were included based on clinical relevance and/or previous demonstration of impact on VI and preventive healthcare use.

### 2.5 Statistical Analysis

In NHATS Round 11, a total of 3,817 participants were sampled, and interview data were obtained by direct contact with either the participant or a proxy respondent. **Figure 1** describes the analytic population. Descriptive statistics were used to characterize the participants by any VI status. Comparisons were performed using Pearson’s chi-squared test for categorical variables and Wilcoxon rank-sum test for continuous variables.

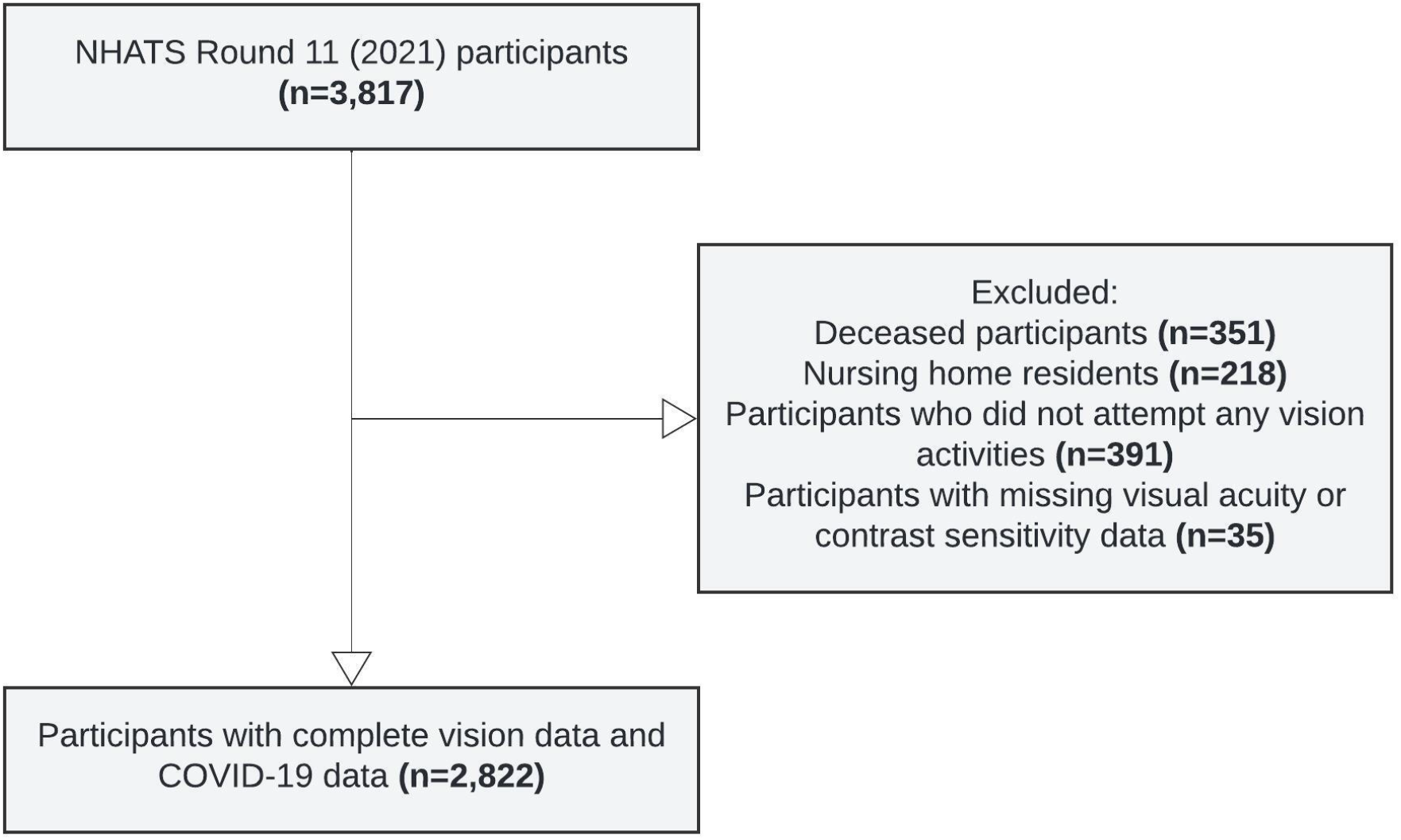

Multivariable logistic regression models were used to investigate the association between various vision measures and COVID-19 vaccination and testing. While our primary exposure was any VI, we constructed separate regression models for each vision variable, including distance VI, near VI, and CSI. We also assessed vision on a continuous scale; distance and near VA (per 0.1 logMAR), and CS (per 0.1 logCS). All analyses accounted for the NHATS complex survey design and models were adjusted for age, sex, race/ethnicity, education, living arrangement, and comorbidities. “Don’t know” and “Refuse” responses were treated as missing values and excluded from the regressions. Statistical significance was defined at p<.05 and 2-sides values are presented. All analyses were performed using Stata/SE 16.1 (StatCorp LLC, College Station, TX, USA) software.

## 3. Results

Among a sample of 2,822 community-dwelling older adults representing 26,182,090 older adults nationally, the majority was female (55%) and White (82%), and 32% had any VI (**Table 1**). Of the total sample, 91% adults were vaccinated for COVID-19, and 53% had ever tested for COVID-19. The prevalence of COVID-19 vaccination did not differ by any VI status, but the prevalence of COVID-19 testing was 11% lower among adults with any VI than adults without VI. Further, COVID-19 vaccination rates were 5% lower in adults with near VI (vs. no near VI), while COVID-19 testing rates were 15% lower in adults with CSI (vs. no CSI). There were no significant differences in the COVID-19 vaccination and testing rates for the other vision measures (**Figure 2**).

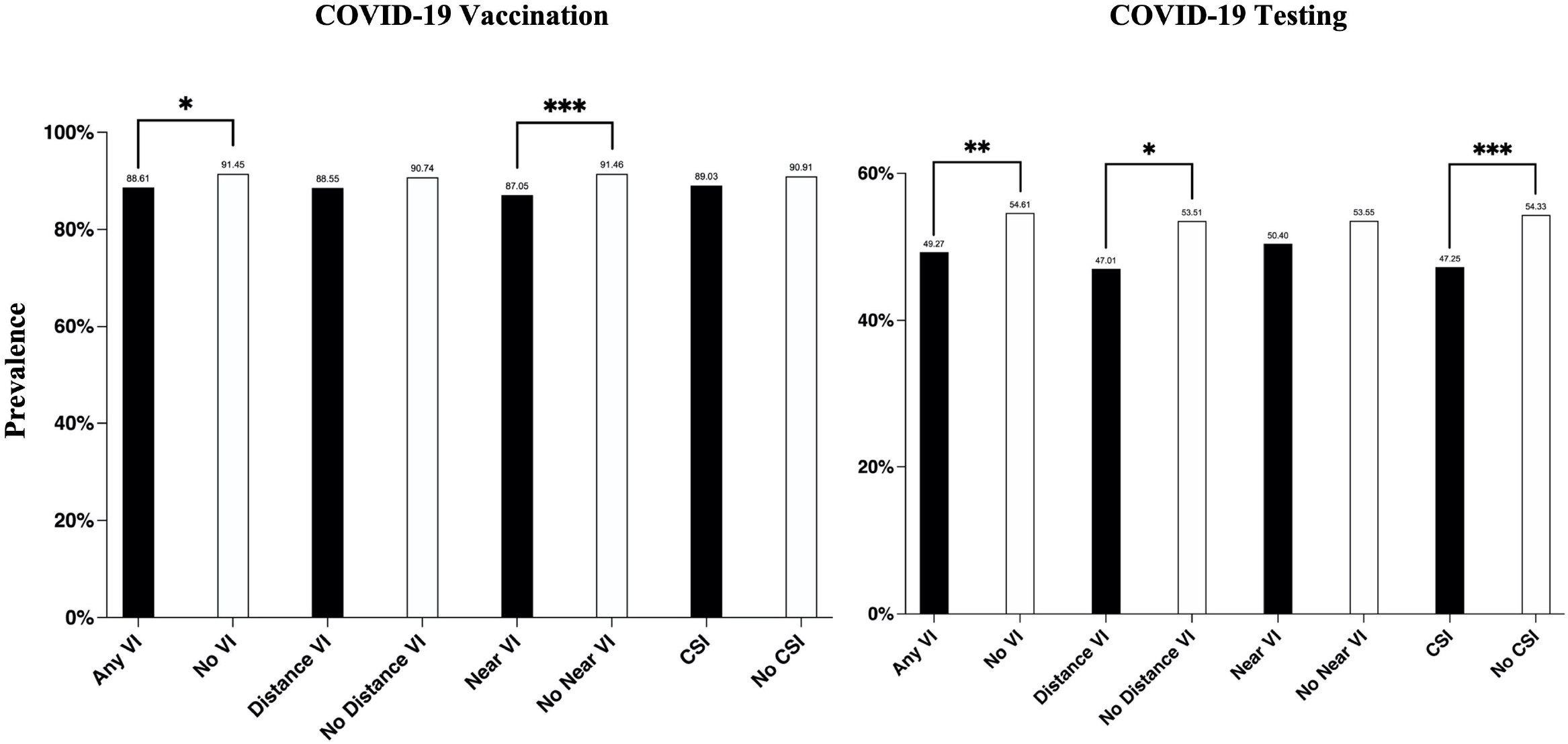

In multivariable logistic regression analysis, older adults with any VI had similar COVID-19 vaccination rates compared to adults without (OR: 0.77, 95% CI: 0.54–1.09), but had lower odds of testing for COVID-19 (OR: 0.82, 95% CI: 0.68–0.97) (**Table 2**). When examining individual objective vision measures categorically, older adults with distance (OR: 0.47, 95% CI: 0.22–0.99) and near (OR: 0.68, 95% CI: 0.47–0.99) VI had lower odds of being vaccinated, and adults with CSI were less likely to test for COVID-19 (OR: 0.76, 95% CI: 0.61–0.95), than their respective peers without impairments. The remaining associations were not significant (p>.05).

When examined on a continuous scale, adults with worse distance (OR: 0.48, 95% CI: 0.24– 0.96), and near (OR: 0.32, 95% CI: 0.16–0.65) VA, (per 0.1 logMAR) were less likely to be vaccinated, however, only adults with worse distance VA were less likely to test for COVID-19 (OR: 0.41, 95% CI: 0.26–0.65). Similarly, adults with worse CS (per 0.1 logCS) were less likely to test for COVID-19 (0.50, 95% CI: 0.36–0.71) but did not have differences in vaccination rates as compared to peers without CSI.

## 4. Discussion

In a nationally representative sample of US older adults, adults with VI were less likely to get vaccinated or tested for COVID-19, depending on the measure of visual function. Interestingly, for each 0.1 logMAR deterioration in distance or near VA, the likelihood of seeking COVID-19 vaccination was reduced to less than half. Similarly, the odds of undergoing COVID-19 testing were equal to or less than half per 0.1 logMAR in distance VA and 0.1 logCS in CS compared to their counterparts without respective impairments. Conversely, near VA (per 0.1 logMAR) trended towards lower odds of undergoing COVID-19 testing, while CS (per 0.1 logCS) trended towards lower odds of vaccination, though neither reached statistical significance.

Our findings are consistent with prior studies showing that people with VI are less likely to utilize COVID-19 healthcare services.^7,22^ This may be due to a range of barriers within the healthcare system, such as issues with accessibility of information and services, and insurance.^4,5,23^ In particular, adults with VI may experience greater difficulties with accessing COVID-19 information on public websites due to poor CS and other webpage accessibility challenges.^4^ Additionally, transportation to drive-through testing sites posed a significant hurdle in accessing testing and vaccination services, which were exacerbated by regulations such as social distancing mandates.^4,5,23^ These findings are concerning, as vaccinations are essential in maintaining health, and preventing severe COVID-19-associated outcomes.^24^ Future work is needed to examine if these disparities in COVID-19 healthcare services translate to worse COVID-19-related health outcomes for older adults with VI.

It is essential to recognize the challenges faced by the VI community and develop strategies to promote health equity and improve access to essential healthcare services. One such strategy is to move towards universal design principles, which aim to create products, environments, and systems that are accessible to everyone, including people with disabilities.^25^ By promoting universal design and addressing inaccessibility, we can ensure that individuals with VI have equal access to healthcare services, thereby improving their independence, health, and overall well-being.

Our study has some limitations. It is unclear what is meant by “been vaccinated for COVID-19?” as NHATS does not specify whether it refers to full primary vaccination or one of two doses. Further, the NHATS does not specify the type of COVID-19 test used (polymerase chain reaction vs. antigen), or testing location (home test vs. or testing center). It is worth noting that some vision measures were associated with lower testing and vaccination, while others were not. This may reflect real-world differences in how various vision measures impact individuals’ functioning but could also be influenced by the sample size and statistical power. Irrespective, overall, these findings suggest that VI is associated with reduced utilization of COVID-19 testing and vaccination services among older adults.

## 5. Conclusion

This study highlights the inequities in the COVID-19 pandemic response for older Americans with vision disability and emphasizes the importance of prioritizing accessibility of healthcare services, especially COVID-19 testing and vaccination services for all people.

## Acknowledgement/Financial Support

None relevant to work presented.

## Data availability statement

Data is publicly available at https://nhats.org/researcher

